# Factors Associated with Active Trachoma among Children in Ebinat District, South Gondar Zone, North West Ethiopia: A community-based cross-sectional study

**DOI:** 10.1101/2022.02.07.22270570

**Authors:** Getalem Aychew Beyene, Nigist Aychew Beyene, Gedefaw Abeje Fekadu

**Author notes:** <u>Email address of Authors</u>. **<u>Corresponding Author</u>**.

## Abstract

**Background:** Despite the availability of effective prevention strategies and treatments, trachoma is still the major cause of infectious blindness worldwide. The majority of blindness from trachoma is currently reported from sub-Saharan Africa, with the greatest burden in Ethiopia. This study aimed to determine the prevalence and factors associated with active trachoma among children aged 1-9 years in Ebinat District.

**Methods:** A community-based cross-sectional study design was conducted in Ebinat District, South Gondar Zone, North West Ethiopia from March 1-30 2018. A multi-stage random cluster sampling technique was employed and all children 1–9 years old from the selected households were clinically examined for trachoma by ophthalmic nurses using the World Health Organization (WHO) simplified clinical grading scheme. A total of 565 children were included. Data were analyzed using SPSS. Multivariable logistic regression was used to identify factors associated with active trachoma among children.

**Result:** The overall prevalence of active trachoma was 36.1% (95% CI: 32.0% - 40.0%). Of all active trachoma cases, 26.5% and 9.6% were trachomatous follicular and trachomatous follicular-intense respectively. Discharge on the eye (AOR=8.23; 95% CI: 4.27-15.87), presence of liquid waste around the main house (AOR=2.31; 95% CI: 1.47-3.61), presence of human feces around the main house (AOR=2.64; 95% CI: 1.61-4.35), unclean faces (AOR= 5.71; 95% CI: 2.18 – 14.96), not washing face and hands frequently (AOR= 2.28; 95% CI: 1.33-3.90), not using soap while washing face and hands (AOR=1.95; 95% CI: 1.21-3.34), and having more flies on children’s face (AOR=1.74; 95% CI: 1.12-2.73) were significantly associated with active trachoma.

**Conclusion and recommendation:** Active trachoma is a major public health problem among children in the Ebinat District. Surgery for trichiasis, antibiotics for active disease, facial hygiene, and environmental improvement (SAFE) strategy at school and community are recommended to lower the burden of trachoma in the Ebinat District, Northwest Ethiopia.

## Background

Trachoma is a contagious eye infection caused by Chlamydia trachomatis, serovars A, B, Ba, or C. (1). Although effective prevention strategies and treatment are available, trachoma still remained the major cause of infectious blindness worldwide (2). Current estimates suggest that 8 million people are visually impaired & 129 million people live in areas known to be trachoma endemic (3-4). Trachoma infections are self-limiting, but repeated infections lead to scarification of the inner eyelid (5).

Trachoma spreads by direct contact with eye and nose discharges from infected individuals or by contact with fomites (6). WHO launched the alliance for the Global Elimination of Trachoma (GET) by 2020 using the ‘SAFE’ (Surgery, Antibiotics, Facial cleanliness, and Environmental Sanitation) strategy (6, 7). The majority of blindness from trachoma is currently reported from sub-Saharan Africa, with the greatest burden in Ethiopia (8, 9, 10).

A study conducted by Yemane showed the prevalence of active trachoma (both Trachomatous Follicular (TF) and Trachomatous follicular-Intense (TI)) children aged 1 - 9 years was 40.14%. The highest prevalence was observed in: - Amhara region (62.6%) followed by, Oromia (41.3%), SNNP (South Nation, Nationalities, and People*’s*) (33.2%), Tigray (26.5%), Somali (22.6%) and Gambella (19.1%) regions (11).

Trachoma is a public health problem in Ethiopia. To reduce the burden of the problem, the Ethiopian government signed the Declaration of Support for VISION 2020. In light of this genuine endeavor of the nation, conducting research to update policymakers about the current status of trachoma is critical. Hence, this study was conducted in Amhara National Regional State, South Gondar Zone, where the highest prevalence of active trachoma was observed in the national survey. Identifying the level and determinants of active trachoma among children are important to strengthen the prevention strategies and achieve the national target. But there are few studies, especially in the study area. Therefore, this study was conducted to update local decision-makers; - regional health bureau and zonal health department programmers on the current status of active trachoma and associated factors among children in the South Gondar zone, Ebinat District.

## Methods

### Study area, period, and population

From March 1 to March 30, 2018, a community-based cross-sectional study was conducted in Ebinat District, South Gondar Zone, Amhara Region, North West Ethiopia, to assess the prevalence and variables associated with active trachoma among children aged 1 to 9 years. Ebinat district is located in the South Gondar Zone, 128 kilometers from Bahir Dar, the region’s seat, and 660 kilometers from Addis Ababa, Ethiopia’s capital, at an altitude of 1730-2500 meters above sea level. The area is classified as a mid-altitude or “Woyna Dega” climatic zone, with annual rainfall ranging from 800-1000 mm. The average yearly and maximum temperatures are around 10 -32°C. According to demographic forecasts from the Federal Democratic Republic of Ethiopia Central Statistical Agency, Ebinat District has a total population of 314,652, with 160,158 men and 154, 494 women (12). A total of 45,934 children aged 1 to 9 years were counted (Ebinat District Health Office Data 2017). Ebinat District comprises 37 kebeles, 2 urban and 35 rural (a kebele is Ethiopia’s smallest administrative unit).

The study’s source populations included all children aged 1 to 9 years old who lived in the Ebinat district. The study’s targets were chosen at random from three kebeles in the district (one urban and two rural). Children from the target kebeles who met the inclusion criteria were included in the study, but those who had an eye injury or were extremely ill, as well as those who were not at home throughout the study time, were excluded.

### Sampling Methods

With a 95% confidence level, a 5% margin of error, a 1.5 design effect, a 62 percent prevalence rate from a prior study (11), and a 10% non-response rate, a minimal sample size of 592 people were enrolled in the study. The study participants were chosen using a multistage sampling process. The study participants were chosen in two steps: first, three kebeles were chosen using a basic random sampling approach, one from an urban kebele and the other two from rural kebeles. Households were chosen at random from each kebele in the second step. The number of houses in each kebele was proportionally distributed based on the number of children in that kebele. If a house had more than one child aged 1 to 9 years, only one was chosen at random to participate in the study. Households without any eligible children were not included in the study. The study only included children who met the inclusion criteria.

### Operational Definition

✓ Clean face: a child who did not have an eye discharge or nasal discharge and/or fly on the face at the time of the visit.
✓ ly density: in the previous studies, household-fly density was determined by examining the presence of flies on children’s faces and around the doorways for about a half minute. Fly density was graded as (1-3 flies), (4-7 flies), and (>7 flies) (13,14).
✓ Active trachoma: a child is considered having active trachoma; if he/she had TF or TI coded “1”; if children have no signs of active trachoma coded “0” for trachoma negative.
✓ Eye examination: each child underwent eye examination by ophthalmic nurses. Each eye was examined with the examiner sitting in front of study participants in the daylight using magnifying binocular lenses (× 2.5) and hand flashlights (torches). The guide used for reporting examination results of the simplified trachoma grading scheme is the one developed by WHO for fieldwork (15). This document briefly and clearly describes the five stages of trachoma.

1. Trachomatous inflammation-(follicles) (TF): five or more follicles, at least 0.5 mm in size, on the “flat” surface of the upper tarsal conjunctiva.
2. Trachomatous inflammation—(intense) (TI): inflammatory thickening of the upper tarsal conjunctiva with more than half of the normal deep tarsal vessels obscured.
3. Trachomatous scarring (TS): scarring of the tarsal conjunctiva (fibrosis).
4. Trachomatous trichiasis (TT): at least one eye-lash rubbing on the eyeball or evidence of eyelash removal.
5. Corneal Opacity (CO): where at least part of the pupil is blurred or obscured (15).

Finally, the presence or absence of each sign of trachoma was recorded on facial observation and eye examination tools for each child.

### Data Collection

Face-to-face interviews, checklist observation, and a pre-tested semi-structured Amharic (the local language) questionnaire were used to collect data from trained four diploma nurses. Four data collectors, two eye examiners, and two field supervisors received two days of training on the study’s objectives, including how to contact respondents, conduct face-to-face interviews, and maintain confidentiality. Following the interview, two trained ophthalmic nurses examined both of the selected child’s eyes using binocular loupe magnifying lenses (2.5x) and hand-flash lights (torches). Eyelashes, cornea, limbus, eversion of the upper lid, and inspection of the tarsal conjunctiva were all evaluated independently for each eye. After each assessment, the required hygienic procedures were done, including wiping hands with alcohol-based hand gel to avoid cross-infection between subsequent participants. For each child, the examination result was recorded on the data collection tool. The WHO-developed fieldwork guide was used to report the findings of the simplified trachoma grading scheme examinations (15).

To check for trachoma, the WHO suggests looking at the eyelids and cornea first for in-turned eyelashes and any corneal opacity. The examiner should then evert the upper eyelid to check the conjunctiva above the tarsal conjunctiva, which is the stiffer region of the upper eyelid. For this purpose, the conjunctiva covering the rounded border of the tarsal plate and the corners of the averted eyelid are not to be checked. Participants who were unavailable throughout the data collecting period were subjected to three checks. The pre-test was conducted outside of the study location in a similar situation, and appropriate modifications were made based on the input. Every day, filed supervisors and main investigators carefully examined the data for accuracy, completeness, and consistency of responses. Double entry of 5% data was made to check for errors.

### Data Analysis

Epidemiological information (EP INFO) version 3.5.2 was used to code, input, and clean data before being exported to statistical packages for social sciences (SPSS) 20 for analysis. To summarize the data, descriptive statistics were employed. A crude odds ratio (COR) was used to determine the degree of relationship between the dependent and predictor variables. To correct for any confounding variables, the adjusted odds ratio (AOR) for each independent variable was obtained using a backward stepwise binary logistic regression analytic model. The multivariable analysis includes variables with a P-value of less than 0.2 in bivariate analysis. When the p-value was less than 0.05, the association was declared statistically significant.

## Results

### Socio-demographic characteristics of study participants

A total of 565 children aged 1 to 9 years were examined for active trachoma in this study, with a 95.4 percent response rate. More than half of the children (53.3%) lived in rural areas. The children’s average age was 4.49 years, with a standard deviation of 2.43 years. Female children made up 51.3 percent of all children. In terms of age, 41% of children are between the ages of one and three years old. Three hundred and fifty-seven heads of households did not have access to formal education, and 242 (42.8%) were farmers. The average family size in the district was 5.4, with a standard deviation of 1.6, and household sizes ranged from three to ten persons. (Table 1).

**Table 1:** Socio-demographic characteristics of participants at Ebinat District, North West Ethiopia, March 2018 (n=565)

| **Factors** | **Frequency** | **Percentage (%)** |
| --- | --- | --- |
| **Child age** |  |  |
| 1-3 years | 231 | 40.9% |
| 4-6 years | 202 | 35.8% |
| 7-9 years | 132 | 23.4% |
| **Sex of the household head** |  |  |
| Male | 420 | 74.3% |
| Female | 145 | 25.7% |
| **Child residence** |  |  |
| Urban | 264 | 46.7 |
| Rural | 301 | 53.3 |
| **Occupation of the household head** |  |  |
| Farmer | 242 | 42.8% |
| Merchant | 131 | 23.2% |
| Government employ | 73 | 12.9% |
| House Wife | 29 | 5.1% |
| Daily Laborers | 37 | 6.5% |
| Others | 53 | 9.5% |
| **Educational status of the household head** |  |  |
| No formal education | 357 | 63.2% |
| Primary | 109 | 19.3% |
| Secondary and above | 99 | 17.5% |

### Water, sanitation, and hygiene status of study participants

About 195 (71.9%) of urban houses obtain drinking water from a water pipe, while 159 (51.5%) of rural households get drinking water from a protected well. The average duration to get water for 71 percent of the families was 30 minutes of walking. Thirty-seven percent of the children came from homes without access to a toilet. Solid and liquid waste disposal pits were found in 62% and 10% of houses, respectively. At the time of the survey, 69% of liquid waste disposal pits were functional. During the survey, liquid waste and human excrement were found in around forty and twenty-three percent of houses, respectively. (Table 2).

**Table 2:** Water, sanitation, and hygiene status of households with children at Ebinat District, North West Ethiopia, March 2018 (n=565).

| **Factors** | **Frequency** | **Percentage (%)** |
| --- | --- | --- |
| **Source of water for consumption** |  |  |
| Pipe water | 232 | 39.6 |
| Protected Well | 223 | 38.7 |
| Protected Spring | 57 | 9.9 |
| River | 49 | 8.4 |
| Other | 19 | 3.4 |
| **Traveling time to a water source** |  |  |
| Less than or equal to 30 minutes | 401 | 71.0 |
| More than 30 minutes | 164 | 29.0 |
| **Solid waste disposal pits available** |  |  |
| Yes | 214 | 37.9 |
| No | 351 | 62.1 |
| **Liquid wastes observed near the main houses** |  |  |
| Yes | 225 | 39.8 |
| No | 340 | 60.2 |

### Face and hands washing practices of study participants

Sixty-three percent of guardians reported they wash their faces and hands once a day for themselves and their children. Fifty-five percent of guardians did not use soap to wash their faces and hands or those of their children. About sixteen percent of the 565 children examined for active trachoma had discharge on their faces, and more than three-quarters had unclean faces. (Table 3).

**Table 3:** Face and hands washing practice of children at Ebinat District, North West Ethiopia, March 2018 (n=565).

| **Factors** | **Frequency** | **Percentage (%)** |
| --- | --- | --- |
| **Frequency of faces and hands washing** |  |  |
| Two or more/day | 208 | 36.8 |
| Once or less/day | 357 | 63.2 |
| **Soap use during faces and hands washing** |  |  |
| Yes | 254 | 45 |
| No | 311 | 55 |
| **Discharge on the faces** |  |  |
| Yes | 90 | 16 |
| No | 475 | 84 |
| **Condition of the faces** |  |  |
| Clean | 105 | 18.5 |
| Unclean | 460 | 81.5 |

### Prevalence of Active Trachoma

The overall prevalence of active trachoma was 36.1% % (95% confidence interval: 32.0% - 40.0%). Of 204 cases with active trachoma, 150 (26.5%) were TF cases while 54 (9.6%) were TI cases. Among all cases of TF, 72 (48%) were girls. The proportion across the ages increases reaching a peak of 36 (17.6%) at the age of 3 years and slowly decreases as age increases. About ninety-one children households headed by farmers in the rural area were affected by active trachoma.

**Figure 1.**
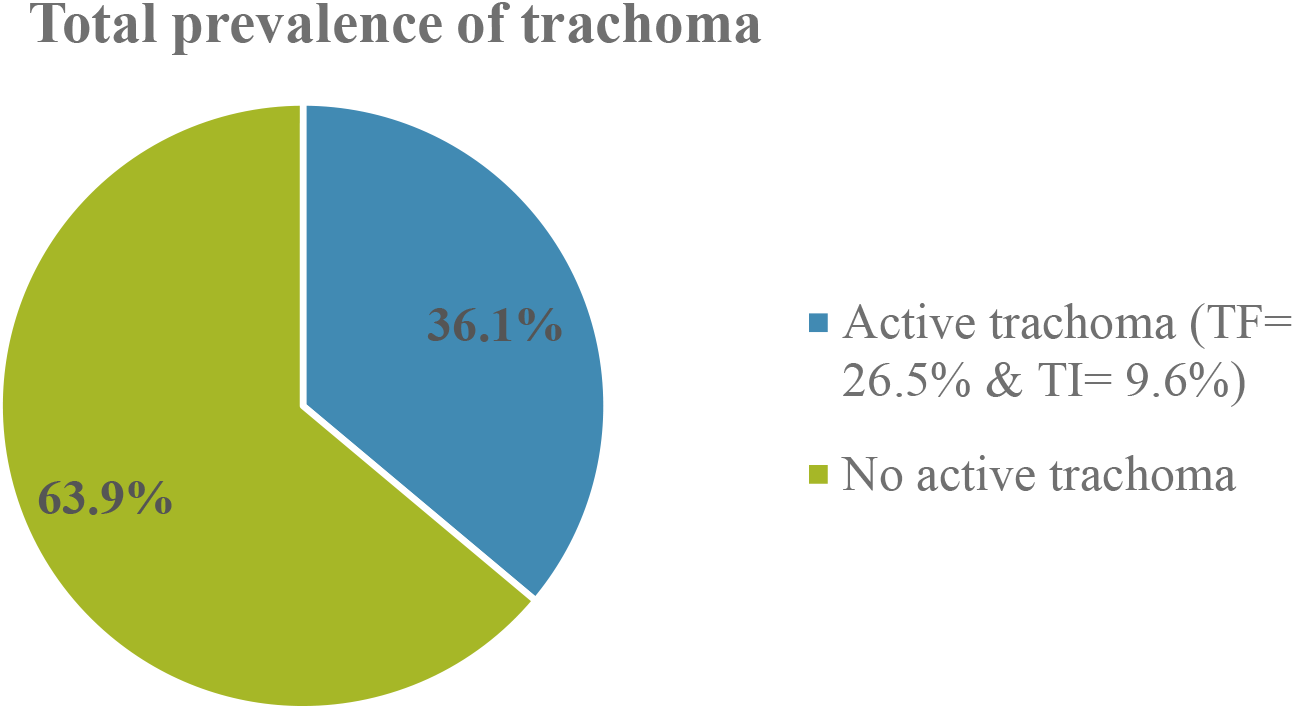
Prevalence of active trachoma in children 1-9 years in Ebinat Woreda from March 1-30, 2018

### Factors associated with active trachoma

In multivariable logistic regression analysis, cleanliness of face, use of soap while washing face and hands, discharge on children’s eyes, presence of liquid wastes, presence of human feces around their main houses, number of flies on children’s faces, and frequency of face washing were significantly associated with active trachoma. The odds of active trachoma among children who had unclean faces were 5.71 times [AOR = 5.71; 95% CI: (2.18– 14.96)] higher than children who had clean faces. Children who didn’t use soap while washing their face and hands were 1.95 times [AOR= 1.95; 95%CI: (1.21–3.34)] more likely to have active trachoma than children who used soaps. The odds of active trachoma among children who had eye discharge had 8.23 times [AOR = 8.23; 95% CI: (4.27– 15.87)] higher compared to children who did not have eye discharge. Children who live in the vicinity which have liquid wastes and human feces around their main houses were 2.31 [AOR= 2.31; 95%CI: (1.47–3.61)] and 2.64 [AOR= 2.64; 95% CI: (1.61 – 3.35)] times more likely to have active trachoma than children who live in the vicinity which have no liquid wastes and feces around their main houses, respectively. The odds of active trachoma among children who had 4 and above flies on their faces were 1.74 times [AOR= 1.74; 95% CI: 1.12-2.73)] higher than children with less than or equal to 3 flies on their faces. Similarly, the odds of active trachoma among children who washed their faces and hands one or less than per day had 2.28 times [AOR = 2.28; 95% CI: (1.33–3.90)] higher compared to those who washed their faces and hands two or more per day. (Table 4).

**Table 4.** Factors associated with active trachoma among children 1 - 9 years in Ebinat District, North West Ethiopia, March 2018 (n=565).

| Variables | Active trachoma |  | Crude OR (95% CI) | Adjusted OR (95% CI) |
| --- | --- | --- | --- | --- |
|  | Yes | No |  |  |
| **Sex** |  |  |  |  |
| Male | 112 | 163 | 1.48 (1.05-2.09)* | 1.05 (0.68-1.63) |
| Female | 92 | 198 | 1 | 1 |
| **Condition of child's face** |  |  |  |  |
| Unclean | 199 | 261 | 15.23 (6.01-38.15)*** | 5.71(2.18-14.96)*** |
| Clean | 5 | 100 | 1 | 1 |
| **Soap use during washing face & hands** |  |  |  |  |
| Yes | 37 | 174 | 1 | 1 |
| No | 167 | 187 | 4.07 (2.70-6.12)*** | 1.95 (1.21-3.34)** |
| **Discharge on eye** |  |  |  |  |
| Yes | 138 | 101 | 16.23 (8.72-30.23)*** | 8.23 (4.27-15.87)*** |
| No | 66 | 260 | 1 | 1 |
| **Liquid waste around the main house** |  |  |  |  |
| Yes | 141 | 131 | 3.93 (2.72-5.67)*** | 2.31 (1.47-3.61)*** |
| No | 63 | 230 | 1 | 1 |
| **Human feces around the main house** |  |  |  |  |
| Yes | 75 | 56 | 4.12 (2.78-6.33)*** | 2.64 (1.61-3.35)*** |
| No | 129 | 305 | 1 | 1 |
| **No of flies on a child's face** |  |  |  |  |
| 0-3 flies | 62 | 206 | 1 | 1 |
| >= 4 flies | 142 | 155 | 3.04 (2.11-4.38)*** | 1.74 (1.12-2.73)** |
| **Frequency of face & hand washing** |  |  |  |  |
| >= 2/day | 34 | 174 | 1 | 1 |
| Once or less/day | 170 | 187 | 4.65 (3.05-7.10)** | 2.28 (1.33-3.90)** |

\*\*\* Significant at p-value $\leq 0.001$ , \*\*Significant at p-value $\leq 0.01$ , \*Significant at p-value $< 0.05$

## Discussion

The prevalence of active trachoma among children aged 1 to 9 years was found to be 36.1% in this study (95 percent CI: 32.0% -40.0%). This prevalence is higher than the WHO’s recommended trachoma eradication target (16). The disparity could be due to a lack of integration of health promotion into basic eye care and a lack of long-term behavioral change to reduce the prevalence of trachoma in the Ebinat District.

The prevalence of active trachoma found in this study was almost same to that seen in earlier studies: Zala District, Gamo Gofa zone (36.7), Ethiopia (36.1%), Uganda (37.3%), Chad (38%), Central African Republic (38%), and Nigeria (37.3%). (17, 18, 19, 20, 21). However, when compared to other studies conducted in the Amhara region (62.6 percent) and Ankober District (53.9 percent), the prevalence of active trachoma was low (22, 23). This could be attributed to geographical disparities, previous studies conducted over several years, low health promotion and practice on personal and environmental hygiene, and insufficient access to health facilities in the studied locations (23).

However, the prevalence is higher than in other studies performed in Ethiopia, such as Kersa District in Jimma Zone (25.2%), Dangla town in Awi Zone (12%), Baso liben District in West Gojam Zone (24.1%), and Dera Woreda in South Gondar Zone (27.9%). (24, 25, 26, 27). This disparity could be due to socio-demographic characteristics of study participants, the study period, low trachoma endemicity, better access to health facilities, and the implementation of a special program integrated into the primary health care system in selected parts of Ethiopia to prevent classical poverty-related diseases such as trachoma (25, 26).

Children who had eye discharge were eight times more likely to have active trachoma as compared to children who had no eye discharge. This finding is in line with two studies from Gambia and a cross-sectional study conducted in Dera woreda, South Gondar Zone, North and South Wollo Zones of Amhara region (27, 28, 29, 30). This finding might be explained by the fact that having discharge on the eye was identified as one of the risk factors for developing active trachoma in the current study. Trachoma is spread from infected to uninfected children’s eyes through discharge from infected children’s hands (fingers), clothing, or eye-seeking flies (31). Furthermore, the Center for International Health identified six Ds in 2007 to aid in memorizing trachoma risk factors. These include dry, dusty, dirty, dung, discharge, and density (overcrowding in the dwelling), all of which could explain the above discovery (32).

Children who washed their faces and hands less than once a day were twice as likely to have active trachoma as those who were cleansed twice or more. This finding was supported by research conducted in the Amhara region’s Gondar Zuriya woreda, the SNNP region’s Dalocha District, and Northern Sudan (33, 34, 35). In contrast to cross-sectional studies conducted in Zala District and North and South Wollo Zones of the Amhara region in 2014 (17, 28), this study found that children who did not use soap when washing their faces and hands were two times more likely to have active trachoma than those who did. Keeping cheeks clean with soap may have aided in avoiding flies’ contact (vector), which is the primary cause and transmission of trachoma. Many studies have shown that keeping one’s face and hands clean with soap throughout the day is crucial in preventing trachoma (25, 31).

Trachoma was found to be strongly linked to a child’s facial cleanliness. Those with dirty faces were six times more likely than children with clean faces to have active trachoma. The current finding is consistent with findings from research in Southern Sudan, Nigeria’s Yobe state, Ankober, the Kersa District of the Jimma Zone, the Baso Liben District of the West Gojjam Zone, Gambia’s Gondar Zuria woreda, and Tanzania’s Gondar Zuria woreda (14, 23, 24, 26, 33, 36, 37, 38). This is because flies are attracted to children who have unclean faces. Eye-seeking flies from children’s faces are more likely to disseminate C. trachomatis-infected ocular secretions among themselves, and a dirty face creates eye discharge that attracts flies (39, 40, 41). The spread of active trachoma by fingers, fomites, or flies would be aided by this infected discharge (37). When comparing children with flies on their faces to their peers, the risks of having trachoma were two times higher. Similar research in Zala District in Southern Ethiopia, Nigeria, Makisegnit Town in Northwest Ethiopia, Brazil, and rural Ethiopia all came up with similar results (17, 21, 38, 42, 43). The involvement of the eye-seeking fly in trachoma transmission, which is still ubiquitous and widespread, could be one explanation.

Children who live in residences with feces around them are three times more likely to develop active trachoma than children who live in households without feces around them, according to the current study. Other research conducted in Dera Woreda and Northern China corroborated this finding (27, 44). Furthermore, children who lived in an area with liquid waste near the main houses were two times more likely than their peers to have active trachoma. This finding was in line with a study conducted in Dera Woreda (27). This could be explained by the fact that the fly, which is the trachoma vector, prefers to breed in human excrement and uncontrolled waste disposal (45).

## Limitations

The Dacron swab for the Chlamydia RNA (Ribonucleic Acid) polymerase chain reaction test was not used to confirm ocular chlamydia infection, which is a study limitation.

## Conclusion

In the Ebinat District, the prevalence of active trachoma among children aged 1 to 9 years surpasses the WHO target for eradication. Active trachoma was identified by discharge on children’s eyes, cleanliness of the face, frequency of face washing, usage of soap while washing face and hands, the number of flies on children’s eyes, and presence of liquid waste around the main residence. To prevent trachoma, it is critical to reinforce and scale up the already existing SAFE strategy, as well as to conduct social behavioral change communication (SBCC) initiatives.

## Data Availability

All relevant data are within the manuscript and its Supporting Information files.

## Abbreviations

AOR: Adjusted Odds Ratio
CI: Confidence Interval
CO: Corneal Opacity
EPINFO: Epidemiological Information
GET: Global Elimination of Trachoma
RNA: Ribonucleic Acid
SAFE: Surgery for trichiasis, Antibiotics for active disease, Facial hygiene, and Environmental improvement
SBCC: Social Behavioral Change Communication
SNNP: South Nation Nationalities of People
SPSS: Statistical Packages for Social Sciences
TF: Trachomatous Inflammation-(Follicles)
TI: Trachomatous Inflammation-(Intense)
TS: Trachomatous Scarring
TT: Trachomatous Trichiasis
WHO: World Health Organization

## Declarations

### Ethical Consideration

This study was approved by the ethical review committee of Haramaya University, College of Health and Medical Sciences, School of Nursing. Permissions letter was also taken from South Gondar Zonal Health Department, Ebinat District Health Office, and selected kebeles. Child assent and informed verbal consent were obtained from the sampled children and their parents. The respondents were also informed that they have the full right to withdraw or refuse at any time from the process. Confidentiality of information given by each respondent was kept properly and anonymity was explained clearly for participants. All children with active trachoma were given two tubes of topical tetracycline eye ointments and instructions on proper use were provided.

## Consent for publication

Not applicable

## Availability of data and materials

Data will be available upon request from the corresponding authors.

## Competing interests

The authors declare that they have no competing interests.

## Funding

No fund was obtained for this study.

## Authors’ contributions

❖ GAB: design, implementation, data analysis and writing the manuscript.
❖ NAB: contributed in conception, continue in design, data analysis.
❖ GAF: contributed in the interpretation of results, writing and editing the manuscript. All authors read and approved the final manuscript.

## Acknowledgments

The authors are grateful to study participants for their kind participation and to all members of the study team. The author’s appreciation was forwarded to Amhara Regional Health Bureau, Ebinat district health office, and health facilities for their technical support and Pastor Assefa and Zelalem Aychew for providing valuable comments. Last but not least our heartfelt appreciation goes to Wizero Abebu Jemberie and Ato Aychew Beyene for their motivation and moral support.

## Notes

### Competing Interest Statement

The authors have declared that no competing interests exist.

### Funding Statement

The author(s) received no specific funding for this work.

## References

1. Pascolini D, Mariotti SPM. Global estimates of visual impairment. Ophthalmology British Journal. 2011.

2. Taylor HR. Trachoma a blinding scourge from the Bronze Age to the twenty-first century, South Yarra, Australia. Haddington Press. 2008.

3. Resnikoff S, Kocur I, Etya’ale DE, Ukety TO. Vision 2020-the right to sight. Ann Trop Med Parasitol. 2008;102(1):3–5.

4. West SK, Munoz B, Mkocha H, Holland MJ, Aguirre A, Solomon AW, et al. Infection with Chlamydia trachomatis after mass treatment of a trachoma hyperendemic community in Tanzania. A longitudinal study. Lancet. 2005;366(1):296–300.

5. World Health Organization. Report of the First Meeting of the WHO Alliance for the Global Elimination of Trachoma. 1997.http://wholibdoc.who.int/hq/1997/WHO/PBL/GET/97.1.pdf. Accessed Decem 18, 2018.

6. Emerson P, Bailey R, Walraven G, Lindsay S. Human and other feces as breeding media of the trachoma vector Musca Sorbens. Med Vet Entomol. 2001;15(3):314–20.

7. World Health Organization. The Trachoma Grading Cards Slide 1/2 SAFE Documents. 2008. Available at: http://www.who.int/blindness/causes/trachoma_documents/en/index.html. Accessed Nov 25 2018.

8. Smith JL, Flueckiger RM, Hooper PJ. The geographical distribution and burden of trachoma in Africa. PLoS Negl Trop Dis. 2013;7(8):1-13. PubMed PMID:e2359; https://doi.org/10.1371/journal.pntd.0002359.

9. Resnikoff S, Pascolini D, Etya’ale D, Kocur I, Pararajasegaram R, Pokharel GP, etal. Global data on visual impairment in the year 2002. Bull World Health Organ. 2004;82(11):844–51.

10. Courtright P, West SK. Contribution of sex-linked biology and gender roles to disparities with trachoma. Emerg Infect Dis. 2004;10(11):2012-16. PubMed PMID: 15550216; PubMed Central PMCID: PMC3328994.

11. Birhan Y, Worku A, Bejiga A, Adamu L, Alemayehu W, Bedri A, et al. Prevalence of Trachoma in Ethiopia. Ethiop.J.Health Dev. 2007;21(3):211–15.

12. Ebenat (woreda) - Wikipedia, the free encyclopedia. 2016. Available from http://en.wikipedia.org/wiki/Ebenat (woreda). Accessed 28 Oct 2018.

13. Ngondi J, Onsarigo A, Matthews F, Reacher M, Brayne C, et al. Effect of 3 years of SAFE (surgery, antibiotics, facial cleanliness, and environmental change) strategy for trachoma control in southern Sudan: a cross-sectional study. Lancet. 2008; 368: 589–595.

14. Emerson PM, Ngondi J. Mass Antibiotic Treatment Alone Does Not Eliminate Ocular Chlamydial Infection. PLoS Negl Trop Dis. 2004:3(3): PubMed PMID: 19333370; PubMed Central PMCID: PMC2657205. e394. doi:10.1371/journal.pntd.0000394.

15. World Health Organization. Primary health care level management of trachoma, Geneva. WHO. 1993.

16. Centre for Disease Control. Guidelines for Management of Trachoma in the Northern Territory, Department of Health and Families. 2008.

17. Mengistu K, Shegaze M, Woldemichael K, Gesesew, H, Markos Y. Prevalence and factors associated with trachoma among children aged 1–9 years in Zala district, Gamo Gofa Zone, Southern Ethiopia. Clinical Ophthalmology. 2016;10:1663–70.

18. Mariotti SP, Pascolini D, Rose-Nussbaumer J. Trachoma: global magnitude of a preventable cause of blindness. British Journal of Ophthalmology. 2009;93(5):563–568.

19. Quicke E, Sillah A, Harding-Esch EM, Last A, Joof H, Makalo P et al. Follicular trachoma and trichiasis prevalence in an urban community in The Gambia, West Africa: is there a need to include urban areas in national trachoma surveillance?”. Tropical Medicine and International Health. 2013;18(11):1344–52. PubMed PMID: 24033501; PubMed Central PMCID: PMC:5405853.

20. Noa Noatina B, Kagmeni G, Mengouo MN, Moungui HC, Tarini A, Zhang Y, et al. Prevalence of Trachoma in the Far North Region of Cameroon: Results of a Survey in 27 Health Districts. PLoS Negl Trop Dis. 2013 May 23;7(5):1-9. PubMed PMID: 23717703; PubMed Central PMCID: PMC:3662655. doi:10.1371/journal.pntd.0002240

21. Mpyet C, Goyol M, Ogoshi C. Personal and environmental risk factors for active trachoma in children in Yobe state, north-eastern Nigeria. Tropical Medicine and International Health. 2010 February;15(2):168–172.

22. Berhane Y, Worku A, Bejiga A. National survey on blindness, low vision and trachoma in Ethiopia -in National Blindness & Low Vision Survey. 2006;1–65.

23. Golovaty I, Jones L, Gelaye B, Tilahun M, Belete H, Kume A, et al. Access to Water Source, Latrine Facilities and Other Risk Factors of Active Trachoma in Ankober, Ethiopia. PLoS ONE. 2009;4(8):1–7: doi:10.1371/journal.pone.0006702.

24. Zerihun, N. Trachoma in Jimma zone, southwestern Ethiopia. Tropical Medicine & International Health. 1997 December;2(12):1115–21.

25. Gedefaw M, Shiferaw A, Alamrew Z, Feleke A, Fentie T, Atnafu K. Current state of active trachoma among elementary school students in the context of ambitious national growth plan: The case of Ethiopia. Health. 2013 October 17;5(11):1768-73. doi.org/10.4236/health.2013.511238.

26. Kassahun K, Tiruneh M, Woldeyohannes D, Muluye D. Active trachoma and associated risk factors among children in Baso Liben District of East Gojjam, Ethiopia. BMC Public Health. 2012 December 22;12:1105. doi:10.1186/1471-2458-12-1105.

27. Alemayehu M, Koye D, Tariku A, Yimam K. Prevalence of active trachoma and its associated factor among Rural and Urban children in Dera woreda, Northwest Ethiopia: A comparative cross-sectional study. Biomed Research International. 2015 March 12;2015:1–8. doi.org/10.1115/2015/570898.

28. Tadesse B, Worku A, Kumie A, Yimer SA. The burden of and risk factors for active trachoma in the North and South Wollo Zones of Amhara Region, Ethiopia: a cross-sectional study. Infectious disease of poverty. 2017;6(143):1–12.

29. E.M. Harding-Escha, T. Edwardsa, A. Sillahb, I. Sarr-Sissohoc, E.A. Aryeec,P. Snellc M.J, et al. Risk factors for active trachoma in The Gambia. Trans R Soc Trop Med Hyg. 2008 December; 102(12):1–13. doi:10.1016/j.trstmh.2008.04.022.

30. Quicke E, Sillah A, Harding-Esch EM, Last A, Joof H, Makalo P et al. Follicular trachoma and trichiasis prevalence in an urban community in The Gambia, West Africa: is there a need to include urban areas in national trachoma surveillance. Trop Med Int Health. 2013 November;18(11):1344–1352. doi:10.1111/tmi.12182.

31. J. Karimurio, M. Gichangi, D. R. Ilako, H. S. Adala, and P. Kilima. Prevalence of trachoma in six districts of Kenya. East African Medical Journal. 2006;83(4):63–68.

32. International Center for Eye Health. London School of Hygiene & Tropical Medicine. 2007, London, UK.

33. Asres M, Endeshaw M, Yeshambaw M. Prevalence and Risk Factors of Active Trachoma among Children in Gondar Zuria District, North Gondar, Ethiopia. Preventive medicine. 2016;1(5):1–8.

34. Alemu Y, Bejiga A. The impact of water supply on trachoma prevalence in Dalocha District, Silte’s Zone, SNNPR. Ethiop Med. 2004; 42: 179–184.

35. Hassan A, Ngondi JM, King JD, Elshafie BE, Ginaid GA, et al. The Prevalence of Blinding Trachoma in the Northern States of Sudan. PLoS Negl Trop. 2011 May; 5(5):1–13. doi:10.1371/journal.pntd.0001027

36. Turner VM, West SK, Munoz B, Katala SJ, Taylor HR, Halsey N, et al. Risk factors for trichiasis in women in Kongwa, Tanzania: a case-control study. Int J Epidemiol. 1993;22:341–7.

37. Ferede AT, Dadi AF, Tariku A, Adane AA. Prevalence and determinants of active trachoma among preschool-aged children in Dembia District, Northwest Ethiopia. Infectious Diseases of Poverty. 2017 Oct 9; 6(128):1–7.

38. Shiferaw D, Moges HG. Risk factors for active trachoma among children aged 1-9 years in Maksegnit town, Gondar Zuria District, Northwest Ethiopia. Saudi J Health Sci. 2013;2(3):202–6.

39. Hsieh YH, Bobo LD, Quinn TO, West SK. Risk factors for trachoma: 6-year follow-up of children aged 1 and 2 years. Am J Epidemiol. 2000;152:204–11.

40. Stocks ME, Ogden S, Haddad D, Addiss DG, McGuire C, Freeman MC. Effect of water, sanitation, and hygiene on the prevention of trachoma: a systematic review and meta-analysis. PLoS Med. 2014;11(2):e1001605.doi:10.1371/journal.pmed.1001605.

41. Polack S, Kuper H, Solomon AW, Massae PA, Abuelo C, Cameron E, et al. The relationship between prevalence of active trachoma, water availability and its use in a Tanzanian village. Trans R Soc Trop Med Hyg. 2006;100:1075–83.

42. Reilly LA, Favacho J, Garcez LM, Courtenay O. Preliminary evidence that synanthropic flies contribute to the transmission of trachoma causing Chlamydia trachomatis in Latin America. Cad. Saúde Pública, 23^rd^ ed. Rio de Janeiro: Brazil; 2007:1682–1688.

43. Cumberland P, Hailu G, Todd J. Active trachoma in children aged three to nine years in rural communities in Ethiopia. Trans R Soc Trop Med Hyg. 2005 February;99(2):120–7.

44. Yumei Z, Xuguang S, Zhiqun W, Ran L, Zhe R. Coming National Program of Epidemiological Survey for Trachoma in China: Prevalence of Trachoma in Northern China. J Clin Exp Ophthalmol. 2013;4(290);1–5.

45. P.M. Emerson,R.L. Bailey, G. E. L. Walraven, and S.W. Lindsay, “Human and other feces as breeding media of the trachoma vector Musca sorbents,” Medical and Veterinary Entomology. 2001;15(3):314–20.

